# Integrating Kolmogorov-Arnold Networks with Ordinary Differential Equations for Efficient, Interpretable and Robust Deep Learning: A Case Study in the Epidemiology of Infectious Diseases

**DOI:** 10.1101/2024.09.23.24314194

**Authors:** Kexin Ma, Xu Lu, Nicola Luigi Bragazzi, Biao Tang

**Affiliations:** School of Mathematics and Statistics, Xi’an Jiaotong University, Xi’an, 710049, People’s Republic of China; The Interdisciplinary Research Center for Mathematics and Life Sciences, Xi’an Jiaotong University, Xi’an, 710049, People’s Republic of China; Institute for Financial Studies and School of Mathematics, Shandong University, Jinan 250100, People’s Republic of China; Department of Clinical Pharmacy, Saarland University, 66123, Saarbrücken, Germany

**Keywords:** KAN, KAN-UDE, Deep Learning, Mechanistic models, Epidemiology

## Abstract

In this study, we extend the universal differential equation (UDE) framework by integrating Kolmogorov-Arnold Network (KAN) with ordinary differential equations (ODEs), herein referred to as KAN-UDE models, to achieve efficient and interpretable deep learning for complex systems. Our case study centers on the epidemiology of emerging infectious diseases. We develop an efficient algorithm to train our proposed KAN-UDE models using time series data generated by traditional SIR models. Compared to the UDE based on multi-layer perceptrons (MLPs), training KAN-UDE models shows significantly improves fitting performance in terms of the accuracy, as evidenced by a rapid and substantial reduction in the loss. Additionally, using KAN, we accurately reconstruct the nonlinear functions represented by neural networks in the KAN-UDE models across four distinct models with varying incidence rates, which is robustness in terms of using a subset of time series data to train the model. This approach enables an interpretable learning process, as KAN-UDE models were reconstructed to fully mechanistic models (RMMs). While KAN-UDE models perform well in short-term prediction when trained on a subset of the data, they exhibit lower robustness and accuracy when real-world data randomness is considered. In contrast, RMMs predict epidemic trends robustly and with high accuracy over much longer time windows (i.e., long-term prediction), as KAN precisely reconstructs the mechanistic functions despite data randomness. This highlights the importance of interpretable learning in reconstructing the mechanistic forms of complex functions. Although our validation focused on the transmission dynamics of emerging infectious diseases, the promising results suggest that KAN-UDEs have broad applicability across various fields.

## Introduction

Dynamical systems, described by ordinary differential equations (ODEs), serve as a cornerstone for modeling processes across a vast array of complex systems. An ODE system, as a mechanistic model, can effectively aid in understanding complex mechanisms by providing a mathematical framework to describe the dynamic behavior of various systems. This allows researchers to simulate, analyze, and predict how different variables interact over time, leading to deeper insights into the underlying processes that govern complex systems in fields such as biology, ecology, engineering, physics, and environmental sciences^1^. In biology, for example, ODEs help model the complex interactions within cellular processes, aiding in drug development understanding of diseases, sch as tumors, such as tumor^2,3^. In the field of public health, they are crucial for simulating and predicting the spread of infectious diseases^4–6^, which informs strategies for the control and prevention of epidemics. In engineering, ODEs model a variety of systems from electrical circuits to mechanical systems, facilitating the design of more efficient and safer technologies^7^. Physics benefits from ODEs through simulations that predict planetary motions and quantum dynamics, while environmental sciences use these equations to model ecological interactions and climate change effects^8^. The versatility and utility of ODEs in these fields stem from their ability to incorporate known laws of nature and principles into models that predict and explain real-world phenomena. This ability makes ODEs indispensable tools in both theoretical research and practical applications, providing insights that are not only profound but also actionable in addressing some of the most pressing challenges.

While mechanistic ODE models excel in scenarios where the underlying mechanism are well-understood and can be accurately quantified, they falter when facing highly nonlinear systems featuring complex interactions that are not easily captured through conventional approaches. Concurrently, the rise of machine learning, particularly deep learning, has equipped us with powerful tools for approximating complex functions and identifying patterns within large datasets. However, these models often lack interpretability and may deviate from established physical laws, rendering them less suitable for scenarios where understanding and adhering to the underlying mechanistic dynamics are essential. Therefore, the methods to merge the power of model-driven and data-driven approaches have become highly appealing. Consequently, the integration of mechanistic models with machine (or deep) learning has emerged as a significant area of interest in recent years^9,10^.

In 2019, Raissi et al. introduced a framework known as Physics-Informed Neural Networks (PINNs)^11^. PINNs incorporate physical laws into the training process as a part of the loss function, addressing both forward and inverse problems involving nonlinear partial differential equations^12,13^. This framework has now been widely adopted across numerous fields as well. Recent studies have extended the concept of PINN to couple neural networks with ordinary differential equation systems^14^, applying them to understand the transmission dynamics of COVID-19 epidemics^15^. In 2020, Rackauckas et al. proposed a framework that embeds neural networks into ordinary differential equations, referred to as Universal Differential Equations (UDEs)^16^. This approach involves neural networks for learning terms with high non-linearity where the mechanisms are unclear, while well-understood mechanisms are modeled using ODEs. UDEs have since been applied in various fields, including estimating the time-varying reproduction number of infectious diseases^17^, which can add new insight in understanding molecular mechanisms^18^. The UDE model offers flexibility by integrating known physical knowledge with data-driven learning, making it suitable for a wide range of complex systems. It is also highly scalable, allowing for customization of both the neural network architecture and the form of the differential equations to fit different systems and problems. However, the combination of neural networks with differential equations can result in a model with numerous parameters and a complex computational process, making the development of an efficient training algorithm for achieving high accuracy both a meaningful and challenging problem. Moreover, in both UDEs and PINNs, the use of neural networks to approximate complex functions or variables still lacks interpretability. That is, while the UDE model demonstrates strong approximation capabilities, the physical interpretability of its outputs--particularly those generated by the neural network--remains challenging. This lack of transparency can hinder understanding of the underlying system dynamics.

The main purpose of this study is to introduce a novel integration of Kolmogorov-Arnold Networks (KANs) with ODEs, aiming to leverage the power of KANs for efficient learning in the coupling of data-driven and model-driven approaches. This approach also utilizes KANs to reconstruct the mechanistic formulation of the learned complex functions, thereby achieving a fully interpretable learning methodology. To this end, we firstly develop the general framework for the hybrid system that combines mechanistic models and KANs. Subsequently, we develop a robust training algorithm that effectively combines the strengths of KANs and ODEs, ensuring efficiency, stability and accuracy in the learning process. Finally, we demonstrate the application of our integrated model framework to several key problems in the field of epidemiology of emerging infectious diseases, showing significant improvements over traditional methods in both predictive accuracy and computational efficiency. The overall research design and corresponding methodology of this study are summarized in Fig. 1.

**Figure 1.**
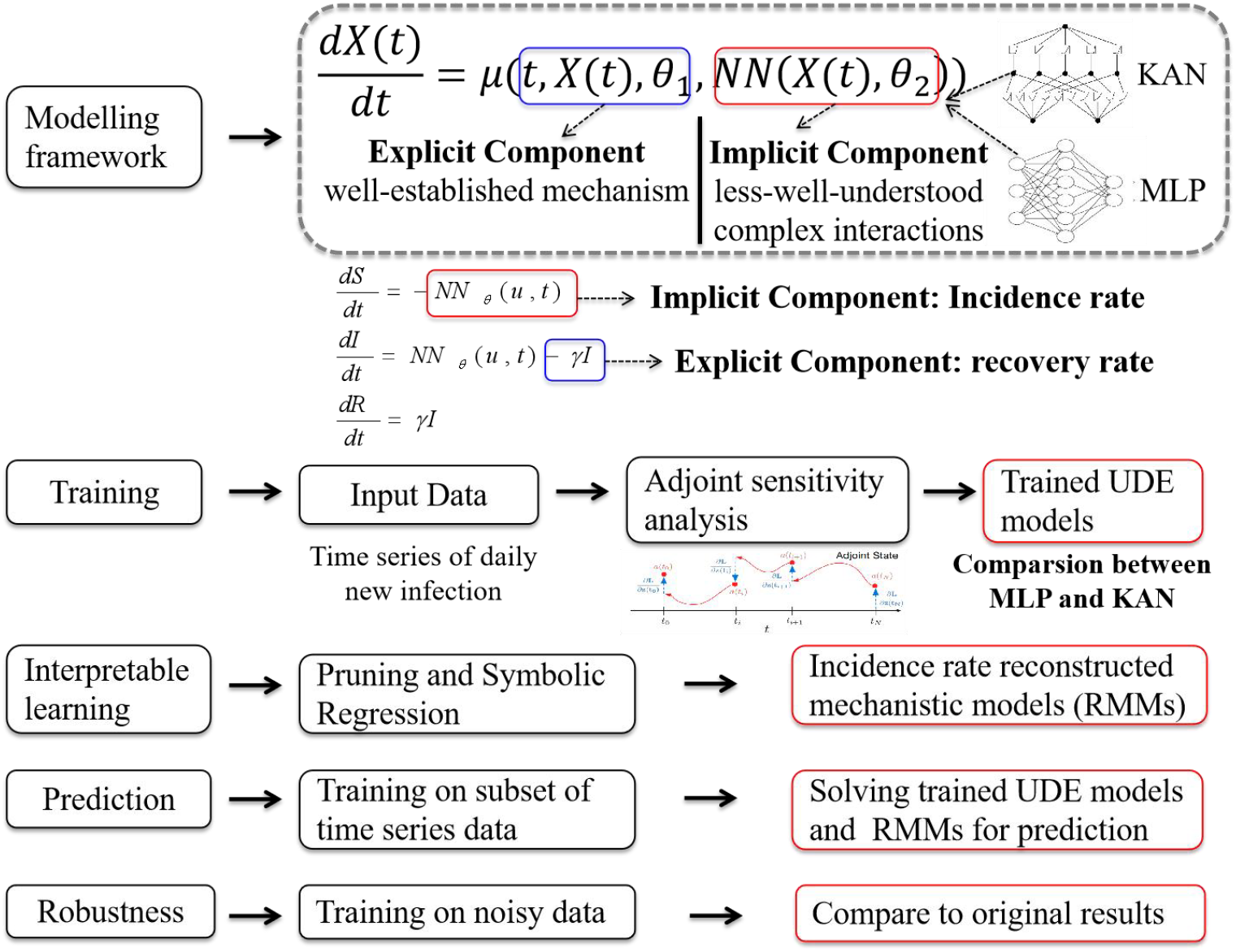
Summary of research design and methodology

## Methods

### Universal Differential Equation (UDE) Models

The Universal Differential Equation (UDE) modelling framework, restricted to the ordinary differential equations, is of the following general form:

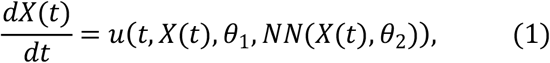

Here, *X*(*t*) is an n-dimensional vector that characterizes the state variables of the complex system at time *t, u* (*t, X* (*t*), *θ* _1_, *NN*(*X* (*t*), *θ*_2_) is the corresponding vector-functions defining the rules governing the changes in those variables. UDE models were developed to address a wide range of dynamic systems by combining neural networks with differential equations, offering a feasible and attractive way to integrate mechanism-driven and data-driven approaches for understanding and predicting complex system behaviors. Specifically, the function *u* consists of two components, as shown in Fig. 1. One component explicitly incorporates the variables *X*, encompassing known physical laws or prior knowledge of the system, which can typically be defined using classical physical principles or experimental data. Correspondingly, *θ*_1_ represents the set of parameters involved in the mechanistic terms. The other component should implicitly involves the state variables but lacks a well-defined mechanistic function. For this reason, we use neural networks to approximate the unknown functions, capturing the complex and hidden interactions within the system. In model (*NN*(*X* (*t*), *θ*_2_)) represents the neural network, *θ*_2_ as the parameters of the network.

Traditionally, the data-driven component in a UDE model is implemented using a multilayer perceptron (MLP). Therefore, the UDE model with an MLP to approximate the unknown functions is expressed as follows:

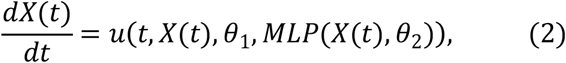

The definitions of the variables and parameters in model (2) are the same as those in model (1). For convenience, we refer to this type of model as an MLP-UDE model.

As noted in the introduction, combining neural networks with differential equations leads to models with numerous parameters and a complex computational process. Therefore, identifying a novel neural network structure to improve training efficiency in UDE models is a key challenge. Kolmogorov-Arnold Networks (KAN) offer an innovative neural network architecture inspired by the Kolmogorov-Arnold representation theorem, providing a practical solution to improve training efficiency and learning accuracy. The representation theorem states that any continuous multivariate function can be expressed as a combination of a finite number of univariate functions and summation operations. Unlike multilayer perceptrons (MLPs), KAN models utilize learnable activation functions along the edges (weights) rather than fixed activation functions at the nodes (neurons). Specifically, the KAN framework replaces traditional linear weight matrices with univariate functions parameterized as splines. Given a continuous multivariate function *f(x)*, it can be represented as:

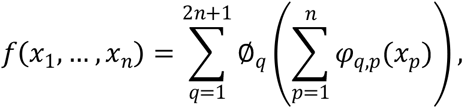

where ∅_*t*_ and *φ*_*q,p*_ are univariate functions. This structure allows KAN model to offer greater flexibility and expressiveness compared to traditional neural network architectures.

In MLP architectures, each layer consists of alternating linear transformations and nonlinear activation functions. In contrast, KAN achieves this through the superposition of function matrices. Each input to a KAN layer is processed through a series of univariate functions, which are then summed up to produce the outputs. KANs are composed of multiple layers, each represented by a function matrix containing univariate functions that map the input to the output dimensions. The computation at each layer can be expressed as:

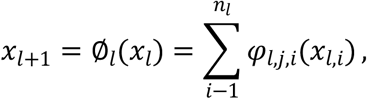

where ∅_*l*_ denotes the function matrix at layer *l*, and *φ*_*l,j,i*_ is the activation function mapping from the *t*th node of layer *l* to the *i*th node of layer *l* + 1. The output of the entire KAN network is the composition of these layers:

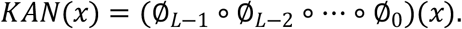

This approach not only avoids the separation of linear and nonlinear components found in MLPs but also allows each weight to be learned as a locally adjustable process based on the input. As a result, KAN exhibits excellent generalization capabilities in high-dimensional spaces, effectively overcoming the curse of dimensionality. In conclusion, MLPs face limitations in scalability when dealing with high-dimensional problems, making it difficult to maintain efficiency while achieving high precision, and they often lack interpretability. Alternatively, the KAN model, with its superior expressive power and computational efficiency in high-dimensional spaces, emerges as an ideal substitute for MLPs in this context.

Therefore, to enhance the expressiveness and interpretability of the UDE model in complex systems, we replace the MLP component in UDE models with KAN, thereby improving the model’s ability to learn unknown nonlinear relationships within the system. This extends the applicability of the UDE framework and leads to the development of a new integrated model, referred to as the KAN-UDE model, which is formulated as follows:

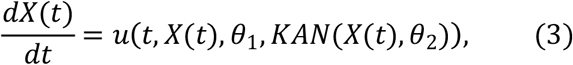

where *KAN*(*X*(*t*),*θ*_2_) represents the unknown rule functions in the complex system as approximated by the Kolmogorov-Arnold Network.

### SIR-type epidemic MLP-UDE and KAN-UDE models

The SIR-type mechanistic compartment model of infectious diseases is of the following form:

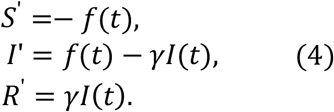

Here, the whole population are divided into three classes according to the epidemic status, i.e. susceptible (*S*), infected (*I*), and recovered (*R*). The parameter *γ* denotes the recovery rate, which can be obtained through the surveillance of the infectious period of the infected individuals. Therefore, it’s a well-established component in building the UDE models for characterizing the population transition from the infectious class to the recovered class. The function *f*(*t*) is the incidence rate of newly infections per unit time. Notably, the incidence rate is a crucial part of epidemic models and often takes various complex forms influenced by multiple factors. For this reason, numerous types of incidence rates have been developed. In this study, four key incidence rates are used, as listed in Table 1. Particularly, for a specific epidemic influenced by various factors, it is often difficult to precisely determine the appropriate incidence rate. Therefore, the incidence rate is treated as the unknown component governing the transition from susceptible to infectious individuals, which will be learned from the data using deep learning.

**Table 1.** MLP-UDE and KAN-UDE models with different incidence rate.

| Incidence rate $f(t)$ | Learned part<br>(parameter values for<br>data simulation) | MLP-UDE models | KAN-UDE models |
| --- | --- | --- | --- |
| Mass action<sup>19</sup> $\beta SI$ | $\beta I$ ($\beta = 0.14$) | $S' = -MLP * S,$<br>$I' = MLP * S - \gamma I(t),$<br>$R' = \gamma I(t).$ | $S' = -KAN * S,$<br>$I' = KAN * S - \gamma I(t),$<br>$R' = \gamma I(t).$ |
| Nonlinear saturated<br>incidence rate<sup>20</sup> $\beta \frac{SI^l}{1+\alpha I^h}$ | $\beta \frac{I^l}{1+\alpha I^h}$<br>($\beta = 0.14, l = 1, h = 1, \alpha = 1$) | $S' = -MLP * S,$<br>$I' = MLP * S - \gamma I(t),$<br>$R' = \gamma I(t).$ | $S' = -KAN * S,$<br>$I' = KAN * S - \gamma I(t),$<br>$R' = \gamma I(t).$ |
| Incidence rate with media<br>effects<sup>21</sup> $\beta e^{-\alpha I} SI$ | $\beta e^{-\alpha I}$<br>($\beta = 0.02, \alpha = 0.5$) | $S' = -MLP * SI,$<br>$I' = MLP * SI - \gamma I(t),$<br>$R' = \gamma I(t).$ | $S' = -KAN * SI,$<br>$I' = KAN * SI - \gamma I(t),$<br>$R' = \gamma I(t).$ |
| Incidence rate with<br>control effects<sup>22</sup><br>$\beta((c_0 - c_b)e^{-rt} + c_b)SI$ | $\beta((c_0 - c_b)e^{-rt} + c_b)$<br>($\beta = 0.014, c_0 = 15, c_b = 10, r = 1$) | $S' = -MLP * SI,$<br>$I' = MLP * SI - \gamma I(t),$<br>$R' = \gamma I(t).$ | $S' = -KAN * SI,$<br>$I' = KAN * SI - \gamma I(t),$<br>$R' = \gamma I(t).$ |

Based on the general frameworks of UDE models, the mechanistic model (i.e., model (4)) can be extended to MLP-UDE or KAN-UDE models by considering part of the incidence rate as the implicit component to be learned from data and the recovery rate as the explicit component, as shown in Fig. 1. The MLP-UDE and KAN-UDE models by setting different parts to be learned related to different incidence rates are listed in Table 1.

Modeling emerging infectious diseases is a crucial tool for understanding transmission mechanisms and predicting epidemics, providing a basis for informed decision-making in disease control. In this study, we focus on applying KAN-UDE models to two key aspects: first, to learn the implicit functions for interpretable deep learning and gain insight into the transmission mechanisms; and second, to achieve accurate and robust predictions of epidemic trends.

### Data

We use the traditional SIR model (i.e. model (4)) to generate the time series data of daily reported case and daily new infections. In detail, the daily new infections at time *t* are calculated by

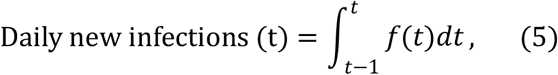

where *f*(*t*) is the corresponding incidence rate in model (4), and four types of incidence rate are used, as listed in Table 1. The daily reported cases at time *t* can be calculated by

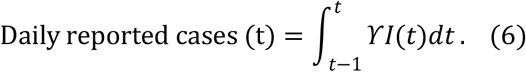

For all the data simulation, we take the population as the density to the whole population, hence the total population is *N* = 1. We assumed that at initial time t=0, the proportion of susceptible individuals is 0.99, the proportion of infected individuals is 0.01, and the recovered population is 0. The parameter values in the SIR model with different types of incidence rate for simulating the time series data are listed in Table 1. The fifth order explicit Runge-Kutta method is used to solve the differential equations. Note that we have generated a separate time series dataset for each model, corresponding to the different incidence rates listed in Table 1. We ran the model (model (4)) from 0 to 99, resulting in a time series dataset of daily new infections (with equation (5)) or daily reported cases (with equation (6)) for each model (i.e. i.e. model (4) with varying incidence rates) over 100 time points. The simulated datasets are shown in Fig. 2. In addition, given the randomness of real data, we re-sampled each time series data (displayed as line chart in Fig. 2) by assuming that the population of the time series at each time point follows a Poisson distribution with a mean of the value of original data^23^. For convenience, we call the time series data with noise as noisy data.

**Figure 2.**
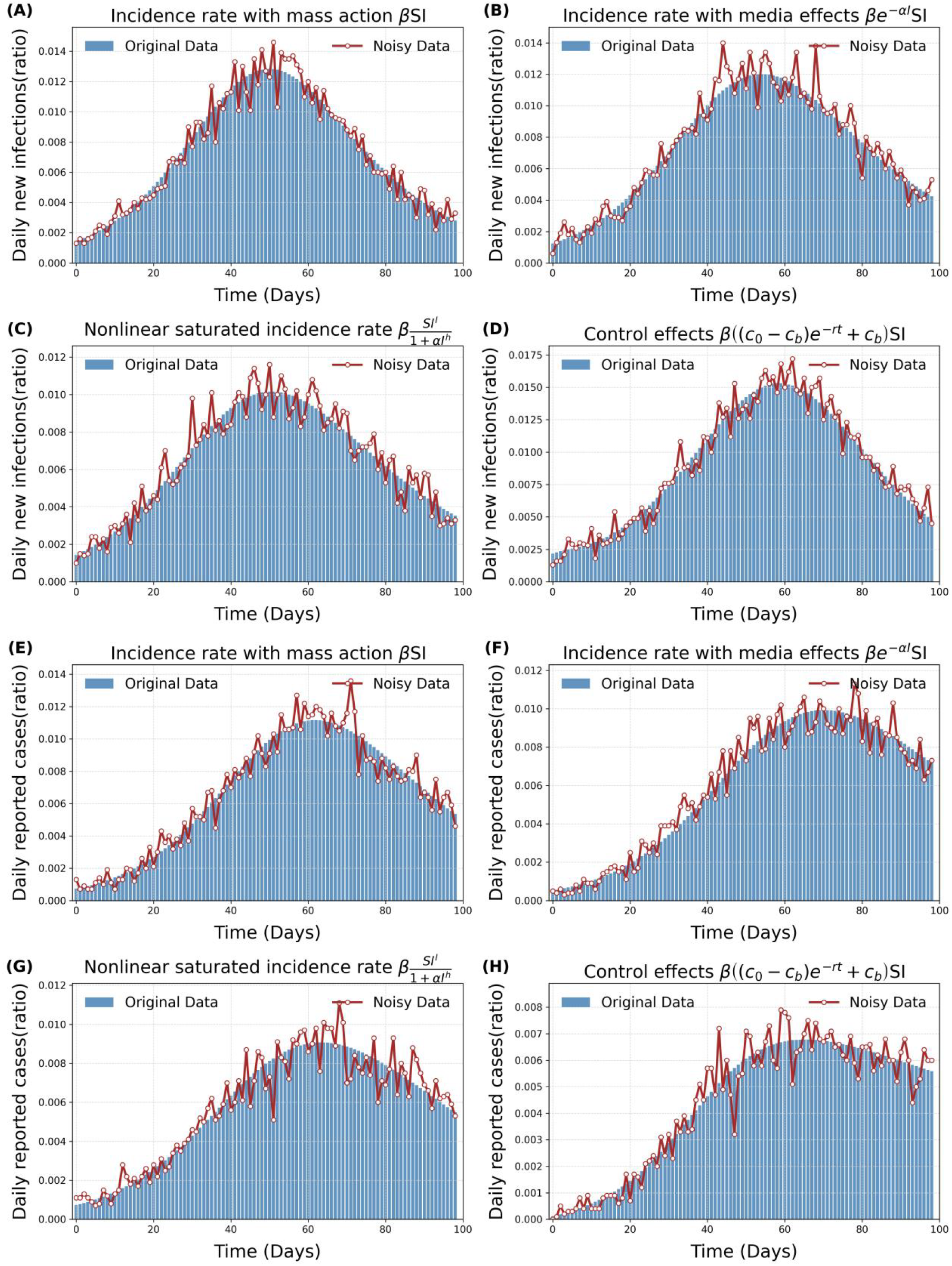
The bar plots represent the simulated time series data of daily new infections (top two rows) and daily reported cases (bottom two rows) generated by the traditional SIR model (i.e., model (4)) with varying incidence rates. For each time series data, the corresponding re-sampled noisy data is depicted as the red line chart.

### Training algorithm

In general, to train the UDE model, we typically begin by fixing the well-known parameter values and initial conditions of the differential equations, informed by prior knowledge. For several emerging infectious diseases, the infectious period is approximately 5 days, allowing us to fix the recovery rate at 1/5. In the case of a newly emerging infectious disease outbreak, the initial recovered population can be set to 0. As we use the simulated data, we also take the initial conditions as the known parameters when training the UDE models.

The next step is to train the model by minimizing the loss function to estimate the unknown parameters in the mechanistic terms and to train the neural networks. The loss function is usually defined as the mean squared error (MSE), given by:

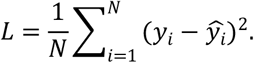

Here, *ŷ*_*i*_ represents the values of the data, *y*_*i*_ is the predicted value by the UDE models, and *t* is the number of data points used for learning. For the epidemics of infectious disease, *ŷ*_*I*_ is the time series data of daily reported cases or daily new infections, which is generated by the mechanistic models in this study.

To proceed the training of the UDE models, adjoint sensitivity analysis is used for calculating the sensitivity of the model output with respect to the parameters. That is, the adjoint equation is defined to compute the sensitivity of the loss function with respect to the model parameters, which is given by:

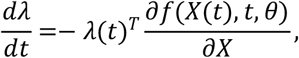

Where *λ* represents the adjoint variable, *X*(*t*) is the state variable, *t* denotes the model parameters, and *f*(*X* (*t*), *t, θ*) represents the rule function governing the changes of the variables. The solution of the adjoint equation involves reverse-time integration, allowing for the efficient computation of gradients that are used for optimizing the model parameters. That is, the gradient of the loss function with respect to each model parameter is computed, which allows for the efficient computation of gradients without the need to directly calculate the partial derivatives for each parameter.

Once the gradients are computed, parameter updates are performed using gradient descent or other optimization algorithms to iteratively reduce the error. After calculating the adjoint variables, the gradient of the loss function with respect to the model parameters is given by:

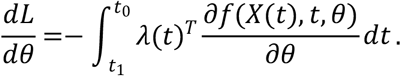

These gradients are utilized to update the parameters in both the ODEs and the neural networks. After calculating the gradients using adjoint sensitivity analysis, an adaptive optimization algorithm, such as the Adam optimizer, is employed to update the model parameters. The Adam optimizer combines momentum and adaptive learning rate adjustments, and its update rule is given by:

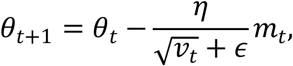

where *θ*_*t*_ represents the current parameter, *t* is the learning rate, *m*_*t*_ is the estimate of the first moment (momentum), and *v*_*t*_ is the estimate of the second moment (variance). The inclusion of momentum and adaptive learning rates in Adam helps in efficiently navigating the parameter space, mitigating issues such as vanishing and exploding gradients.

With the above preparation, the training process begins with model initialization. For the ordinary differential equation (ODE) component, parameter initialization is based on known physical principles, ensuring that the model parameters are physically meaningful. For the knowledge-enhanced neural networks (KANs) component, the initial weights are generated from the control points of the spline functions, ensuring that the model starts with sufficient expressive power and favorable convergence properties. The initial weights are randomly initialized using either a uniform or normal distribution. The positions of the spline function control points and the initial weight values are carefully designed to enable the network to effectively learn complex nonlinear relationships.

Beside the above design of the training process, we also adapt a series of algorithms to facilitate the training and optimizing efficient of the KAN-UDEs model, for enhancing computational efficiency, and improving predictive accuracy. This includes

1. Unlike the original KAN network, which expands all intermediate variables to apply different activation functions, we use a method that activates inputs with various basis functions and then linearly combines them. This recalculation approach significantly reduces memory costs by simplifying computations to basic matrix multiplications, and it naturally supports both forward and backward propagation.
2. During training, the KAN-UDE models adaptively adjusts the density of the spline grid to more accurately capture complex system behaviors. Specifically, in regions with higher errors, a finer grid is used to capture more detailed features, while in regions with lower errors, a coarser grid is maintained to conserve computational resources. This dynamic adjustment strategy ensures the model’s efficiency and accuracy.

### Function reconstruction

To enhance the interpretability and practical applicability of the UDE models, we perform pruning process and symbolic regression on the functional relationships identified by the KAN network, to derive explicit mathematical descriptions of the mechanisms in the implicit component, i.e. to reconstruct the functions in the incidence rate in this study. Initially, the pruning process begins by introducing L1 regularization and entropy regularization to sparsify the activation functions, thereby reducing redundant connections within the networks. Here, L1 regularization measures the average output magnitude of the activation functions, while entropy regularization controls the smoothness of the activation function distribution to further enhance sparsity. By defining L1 norms and entropy metrics on the input and output nodes, the pruning strategy ensures that only nodes critical to the network’s predictions are retained, significantly reducing the network’s complexity. This process removes non-essential nodes from the network structure by controlling a threshold.

We then conduct the symbolic regression to reconstruct the functions based on the reduced KAN by pruning process. Taking the outputs of the trained KAN in the UDE models as the data information for function reconstruction, we initially search a best-fit type of mechanistic functions, such as sin, log, exp, of the activation functions of each connection between two nodes in the network. We therefore obtain a general form of the functions with several affine transformation parameters in each activation function. We further use grid search and linear regression to estimate the affine parameters by minimizing the residual sum of squares between the output of the reduced KAN and the predicted values of the learned functions. As a result, the implicit interactions in the complex system are of explicit mathematical expression, and the UDE models become fully mechanistic models. We call the KAN-UDE model with reconstructed forms of the functions in the implicit component as reconstructed mechanistic model (RMM) for convenience.

### Epidemic predictions

We use the subset of the time series data to train the KAN-UDE models, and estimate parameters and learn neural networks. We considered three scenarios in terms of the data set: a subset including the data from initial to a time point far before the peak time, or to a time point near the peak time, or to a time point after the peak time. In parallel, we use the same subset of the time series date to reconstruct the functions of the incidence rate to obtain the reconstructed mechanistic models (RMMs). Then, we can solve the trained KAN-UDE models or the RMMs to predict the epidemic trends, here fifth order explicit Runge-Kutta method is used to solve the model in terms of daily new infections or daily reported cases. Furthermore, the rest of the time series data are used to test the prediction accuracy of different models.

### Robustness analysis

Given the robustness of the results, we primarily use noisy time series data to train the model, learn the neural networks, reconstruct the incidence rate functions, and predict the epidemic trends. We then conduct a thorough comparison between the results obtained from the original data and the noisy data. It is worth noting that using a subset of the time series data to train the model, reconstruct the functions, and predict epidemic trends also serves as a key aspect in testing the robustness of our models’ results.

## Main results

### Training results

Using the time series data of daily new infections, we independently trained all the MLP-UDE and KAN-UDE models listed in Table 2 by selecting various neural network configurations. We firstly showed the results related to the incidence rate of mass action in Fig. 3, that is, the results using the time series data, generated from the mechanistic model with a incidence rate of mass action, to train the MLP-UDE and KAN-UDE models. It is evident from Fig. 3 that the KAN-UDE model achieves significantly higher accuracy across all neural network configurations after 5,000 iterations, indicating a superior fit to the time series data. Specifically, using the learning accuracy defined in Supplementary Information, the KAN-UDE models achieved 100% accuracy across all configurations except 2-5-1 after 5,000 iterations, as shown in Table 2. In contrast, the MLP-UDE model only reached 100% accuracy with the 2-64-1 configuration. Furthermore, with simpler configurations such as 2-8-1 and 2-5-1, the accuracy of the MLP-UDE model was only 8.33% and 13.3%, respectively. The declining trend in training accuracy with a simpler network configuration for MLP-UDE models is intuitively visible in Fig. 4. However, the KAN-UDE model still achieved 100% accuracy with the 2-8-1 configuration, indicating greater flexibility in learning across different configurations.

**Table 2.** Comparison of training process and training outcomes between MLP-UDE and KAN-UDE models across various configurations.

| Incidence rate used for generating the time series data | Network Configuration | Accuracy (%) |  | Time (s) |  | Iterations to 100% Accuracy |  | Loss (after 5000 iterations) |  |
| --- | --- | --- | --- | --- | --- | --- | --- | --- | --- |
|  |  | MLP | KAN | MLP | KAN | MLP | KAN | MLP | KAN |
| Mass action | 2-128-1 | 95 | 100 | 628 | 1242 | - | 1100 | $2.38 \times 10^{-5}$ | $2.20 \times 10^{-6}$ |
| | 2-64-1 | 100 | 100 | 670 | 1264 | 4600 | 1800 | $6.45 \times 10^{-4}$ | $8.59 \times 10^{-7}$ |
| | 2-32-1 | 66.67 | 100 | 711 | 1263 | - | 3900 | $1.02 \times 10^{-4}$ | $2.28 \times 10^{-6}$ |
| | 2-16-1 | 73.33 | 100 | 732 | 1178 | - | 1900 | $1.04 \times 10^{-4}$ | $7.69 \times 10^{-7}$ |
| | 2-8-1 | 28.33 | 100 | 645 | 1311 | - | 4900 | $8.17 \times 10^{-4}$ | $3.72 \times 10^{-5}$ |
| | 2-5-1 | 8.3 | 80 | 488 | 1201 | - | - | $1.36 \times 10^{-4}$ | $4.99 \times 10^{-5}$ |
| Nonlinear saturated incidence rate | 2-16-1 | 94.17 | 100 | 706 | 1389 | - | 3600 | $3.67 \times 10^{-5}$ | $2.45 \times 10^{-6}$ |
| Incidence rate with media effects | 2-16-1 | 65 | 100 | 693 | 1130 | - | 3200 | $1.11 \times 10^{-4}$ | $1.38 \times 10^{-5}$ |
| Incidence rate with control effects | 2-16-1 | 83.33 | 100 | 732 | 1051 | - | 4300 | $4.52 \times 10^{-5}$ | $1.04 \times 10^{-5}$ |

Note that, “-” means that the model did not reach 100% accuracy after 5,000 iterations.

**Figure 3.**
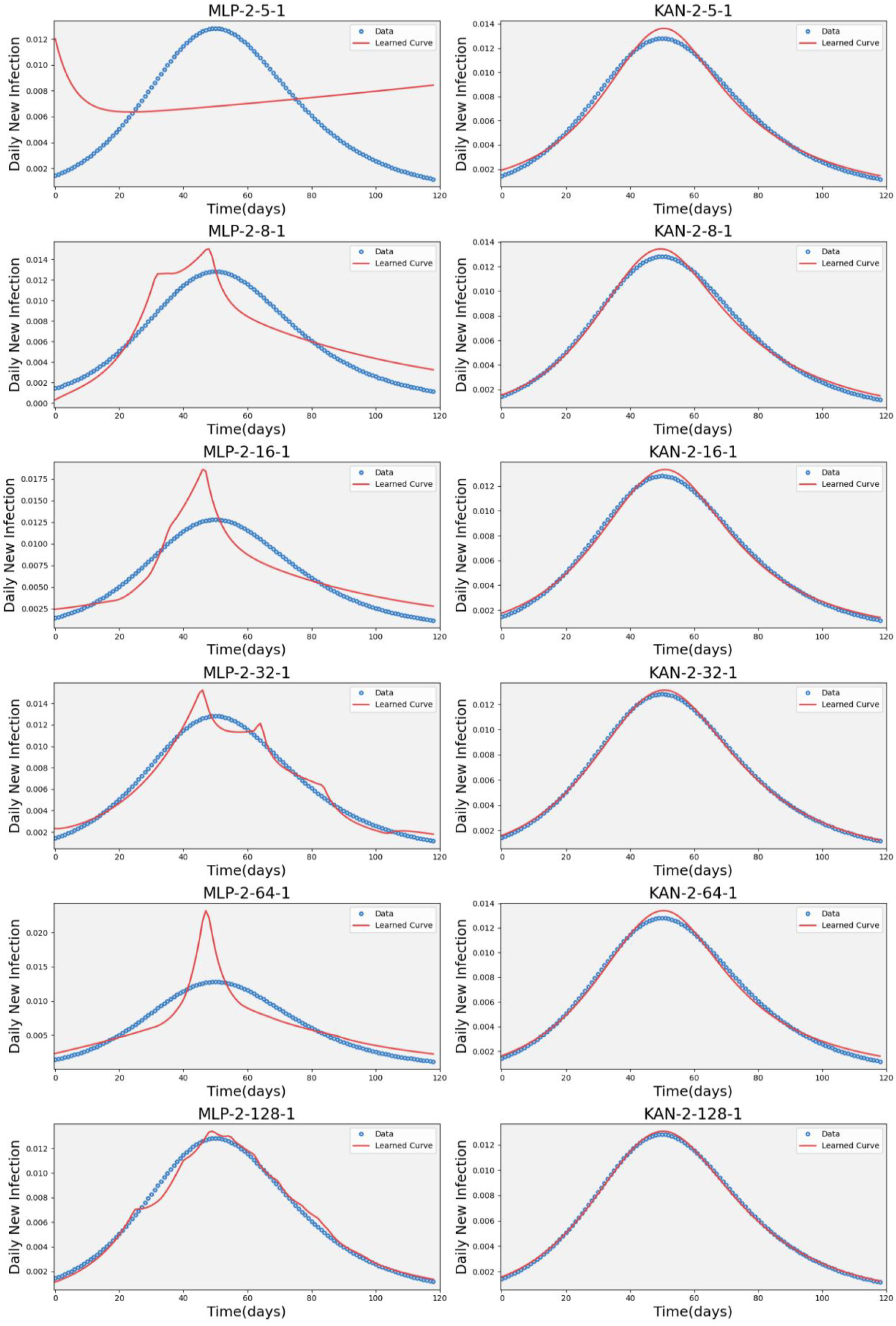
Training results after 5000 iterations for MLP-UDE and KAN-UDE models across various configurations, where the left panel is for MLP-UDE model while the right panel for KAN-UDE model. Here the time series data of daily new infections generated by the traditional SIR model with the incidence rate of mass action is used.

**Figure 4.**
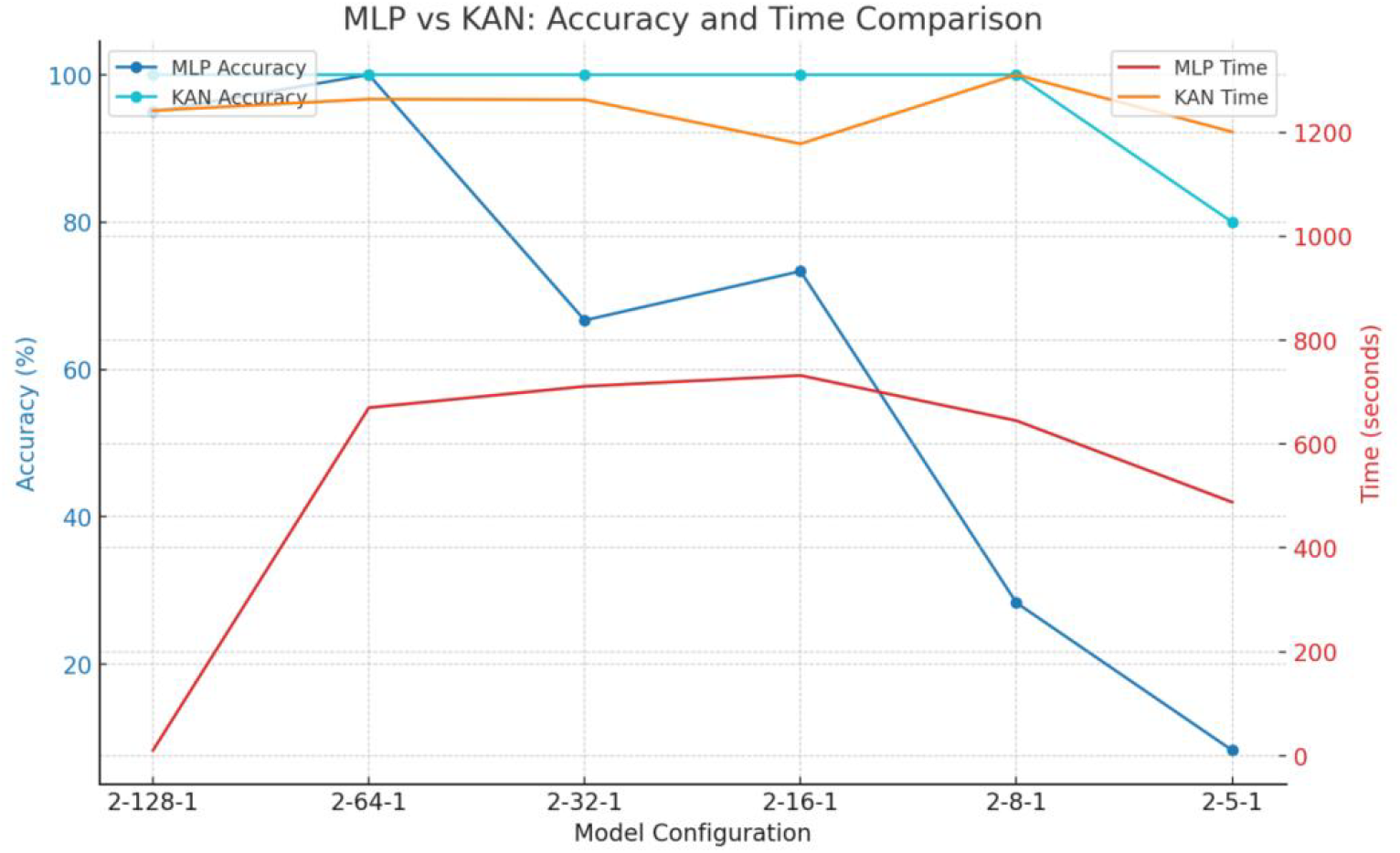
Accuracy and training time for both MLP-UDE and KAN-UDE models across different configurations.

On the other aspect, we also counted the number of iterations needed to achieve 100% accuracy for the two types of UDE models, as detailed in Table 2. From Table 2, it is evident that the KAN-UDE model requires significantly fewer iterations to reach 100% accuracy compared to the MLP-UDE models. For example, with a 2-128-1 configuration, the KAN-UDE model achieved 100% accuracy within 1,400 iterations, whereas the MLP-UDE model required 4,800 iterations to reach the same accuracy. The conclusion that the KAN-UDE model outperforms the MLP-UDE model when comparing the loss of the two UDE models after 5,000 iterations, as shown in Table 2.

Overall, we find that the KAN-UDE modesl outperform the MLP-UDE models across multiple metrics. When using the time series data generated from the traditional SIR model with the other three types of incidence rates to train the UDE models, we observe the similar results in terms of learning accuracy and final loss when comparing the MLP-UDE models and the KAN-UDE models, as shown in Fig. 5 and Table 2. Therefore, the KAN algorithm proves to be more effective and efficient in scenarios where high accuracy and low loss are critical, despite requiring longer computation time per iteration. Furthermore, using the estimated parameters and trained neural networks, we solved for the variables (i.e. *S*(*t*), *I*(*t*), *R*(*t*)) in the KAN-UDE models, as shown in SI Fig. 1. SI Fig. 1 demonstrates that the simulated curves of all variables align closely with the simulated data. This indicates that training the KAN-UDE models with a single time series dataset can accurately capture the behavior of all the variables.

**Figure 5.**
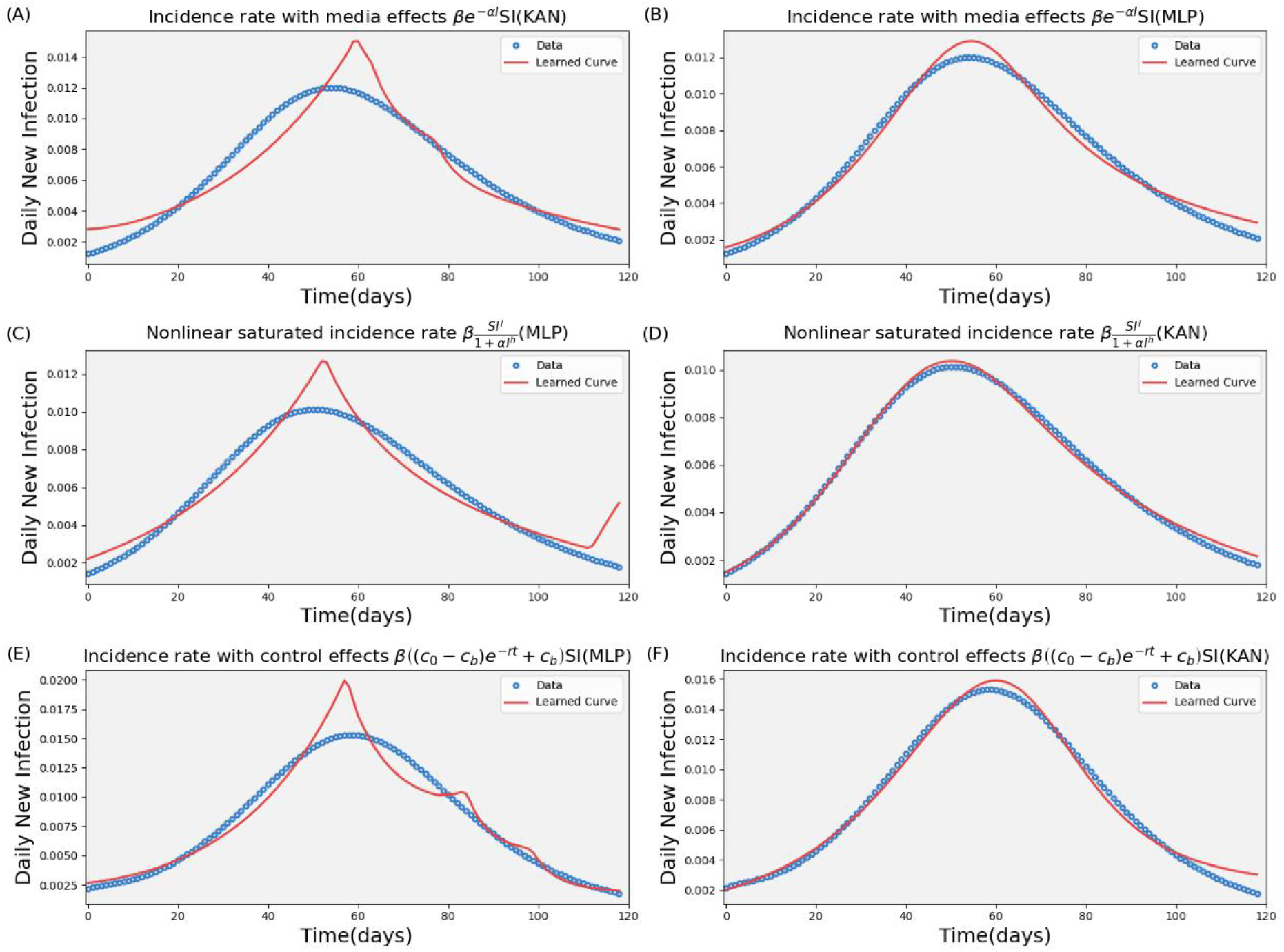
Training results after 5000 iterations for MLP-UDE and KAN-UDE models. Here, we use the time series data generated from the traditional SIR model with three different types of incidence rate, as marked in the title of each sub-figure. The network configuration is fixed as 2-16-1.

### Interpretable learning

To verify the interpretability of the KAN model, we reconstructed the implicit functions of the incidence rate after the training process. The reconstruction results are detailed in Table 3. As shown in Table 3, the KAN model accurately reconstructed all the incidence rate functions across various types, perfectly matching the original settings of the functions used to generate the time series data. Notably, after reconstructing the incidence rate functions, the implicit components of the KAN-UDE models gained explicit mechanistic meaning. As a result, the KAN-UDE model now becomes a fully mechanistic model, referred to as a reconstructed mechanistic model (RMM). This demonstrates the KAN-UDE model’s strong capability to represent the functional forms of these models, offering superior generalization and enhanced interpretability.

**Table 3.** Performance comparison of KAN-UDE models with different incidence functions.

| Incidence rate used for generating the time series data | Time(s) | Iteration | Loss | Reconstructed functions |
| --- | --- | --- | --- | --- |
| $0.4I$ ($\beta SI$) | 427s | 1200 | $5.51 \cdot 10^{-6}$ | $0.4I$ |
| $\frac{I}{1+I} \left( \beta \frac{SI^l}{1 + \alpha I^h} \right)$ | 385s | 1400 | $3.12 \cdot 10^{-5}$ | $1 - \frac{I}{1+I}$ |
| $\frac{I}{1+2I} \left( \beta \frac{SI^l}{1+\alpha I^h} \right)$ | 479s | 1800 | $1.15 \cdot 10^{-5}$ | $0.5 - \frac{0.05I}{0.2+0.1I}$ |
| $\frac{I}{1+5I} \left( \beta \frac{SI^l}{1+\alpha I^h} \right)$ | 393s | 1600 | $2.20 \cdot 10^{-5}$ | $0.5 - \frac{0.05I}{0.2+0.1I}$ |
| $e^{-0.5I} (\beta e^{-\alpha I} SI)$ | 533s | 1000 | $3.12 \cdot 10^{-6}$ | $e^{-0.5I}$ |
| $e^{-I} (\beta e^{-\alpha I} SI)$ | 397s | - | $1.21 \cdot 10^{-4}$ | $e^{-I}$ |
| $5.0 + 10e^{-0.2t}$<br>$\beta((c_0 - c_b)e^{-rt} + c_b)SI$ | 624s | - | $2.2 \cdot 10^{-5}$ | $5.0 + 10e^{-0.2t}$ |

### Epidemic predictions

We employed KAN-UDE models, MLP-UDE models, and RMMs to predict epidemic trends by training the models on a subset of the time series data and reconstructing the incidence rate functions, as shown in Figs. 6-8. As demonstrated in Figs. 6-8, the RMMs consistently provide perfect predictions of the epidemic trends as the red dashed curves perfect matches the testing data, because KAN precisely reconstructs the incidence rate functions. This holds true whether the subset of time series data is taken from the initial period up to 40 days before the peak, around 60 days near the peak, or 80 days after the peak. In contrast, the predictions by the KAN-UDE models and MLP-UDE models are significantly less accurate than those of the RMMs, showing poorer robustness due to the lack of a mechanistic form for the incidence rate. A closer examination reveals that the KAN-UDE model also outperforms the MLP-UDE model in terms of the prediction accuracy, because of achieving higher learning accuracy after the same number of iterations during training. In particular, the KAN-UDE model performs well in short-term predictions (7-day forecasts) for several scenarios (comparing the green dashed curves with the testing data in Fig. 7(B) and Fig. 8(A)), especially when trained on longer time series data, as evidenced by comparing Fig. 8 to Fig. 6. However, prediction accuracy significantly decreases in long-term forecasts, as indicated by the increasing error between the test data and the model’s predicted values.

**Figure 6.**
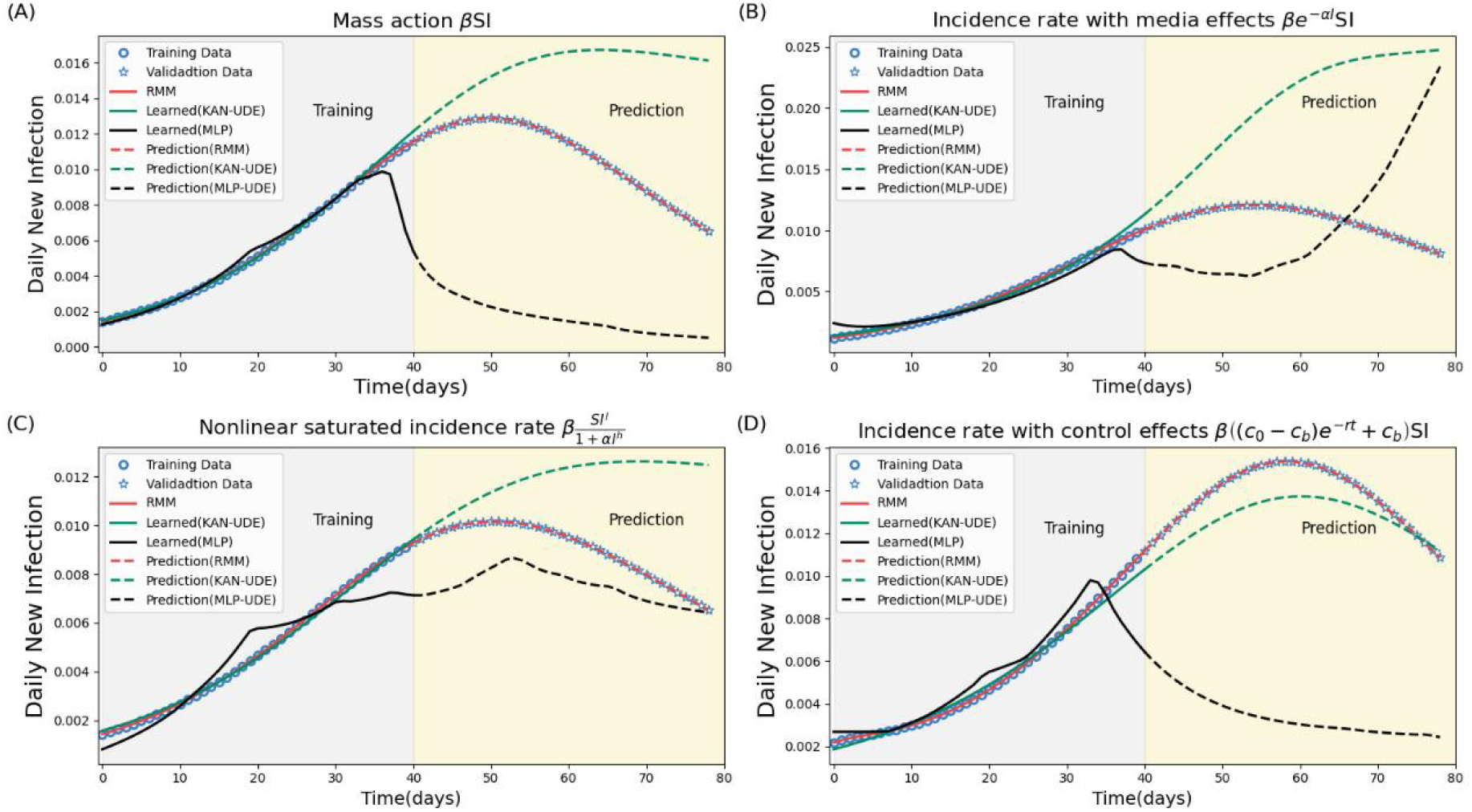
Epidemic prediction by KAN-UDE model, MLP-UDE models, and RMM by training the model with a subset of the time series data. Here the data from original period up to 40 day is used for training and the remaining data (validation data) for testing the prediction accuracy. The training process is stopped after 5,000 iterations.

**Figure 7.**
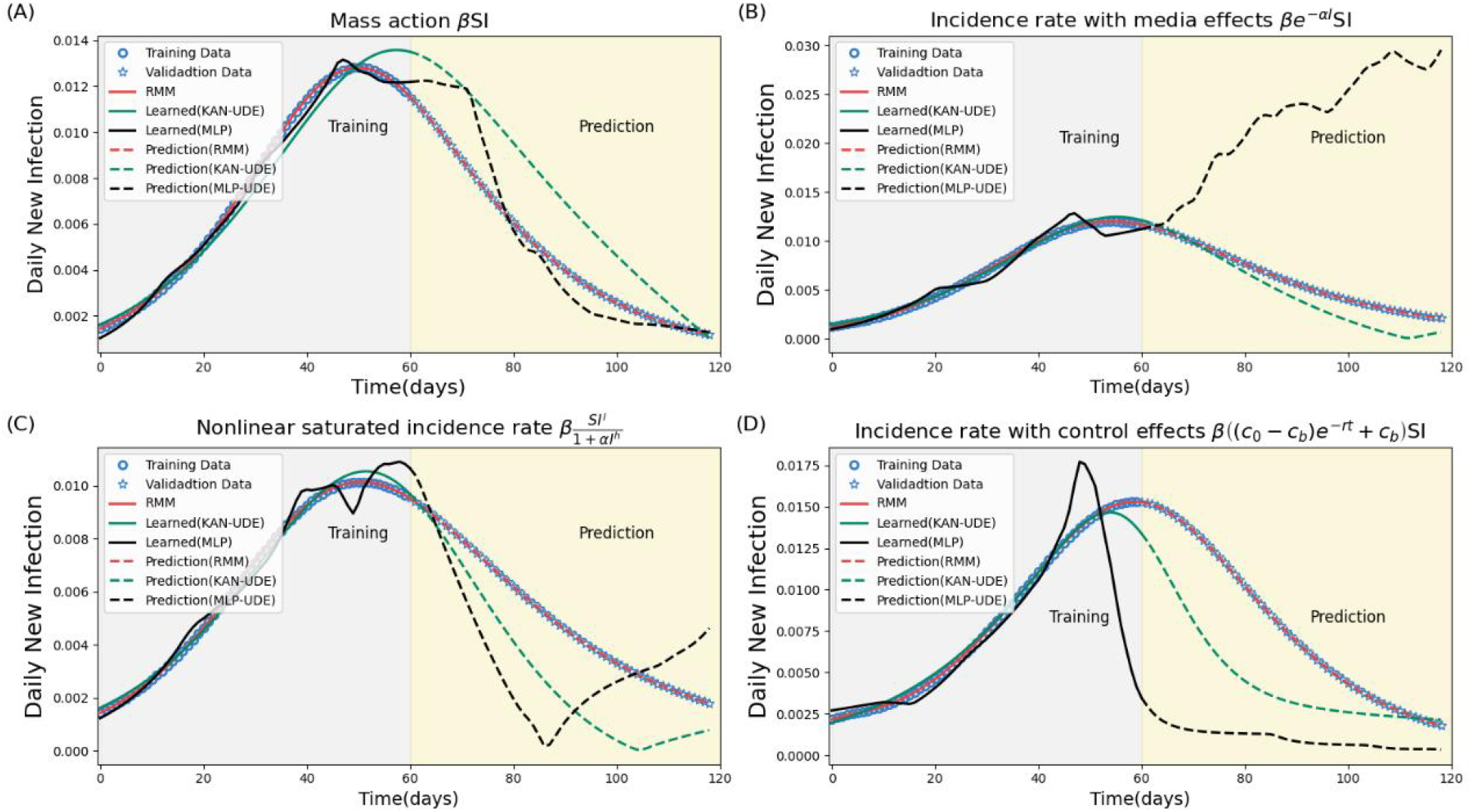
Epidemic prediction by KAN-UDE model, MLP-UDE models, and RMM by training the model with a subset of the time series data. Here the data from original period up to 60 day is used for training and the remaining data (validation data) for testing the prediction accuracy. The training process is stopped after 5,000 iterations.

**Figure 8.**
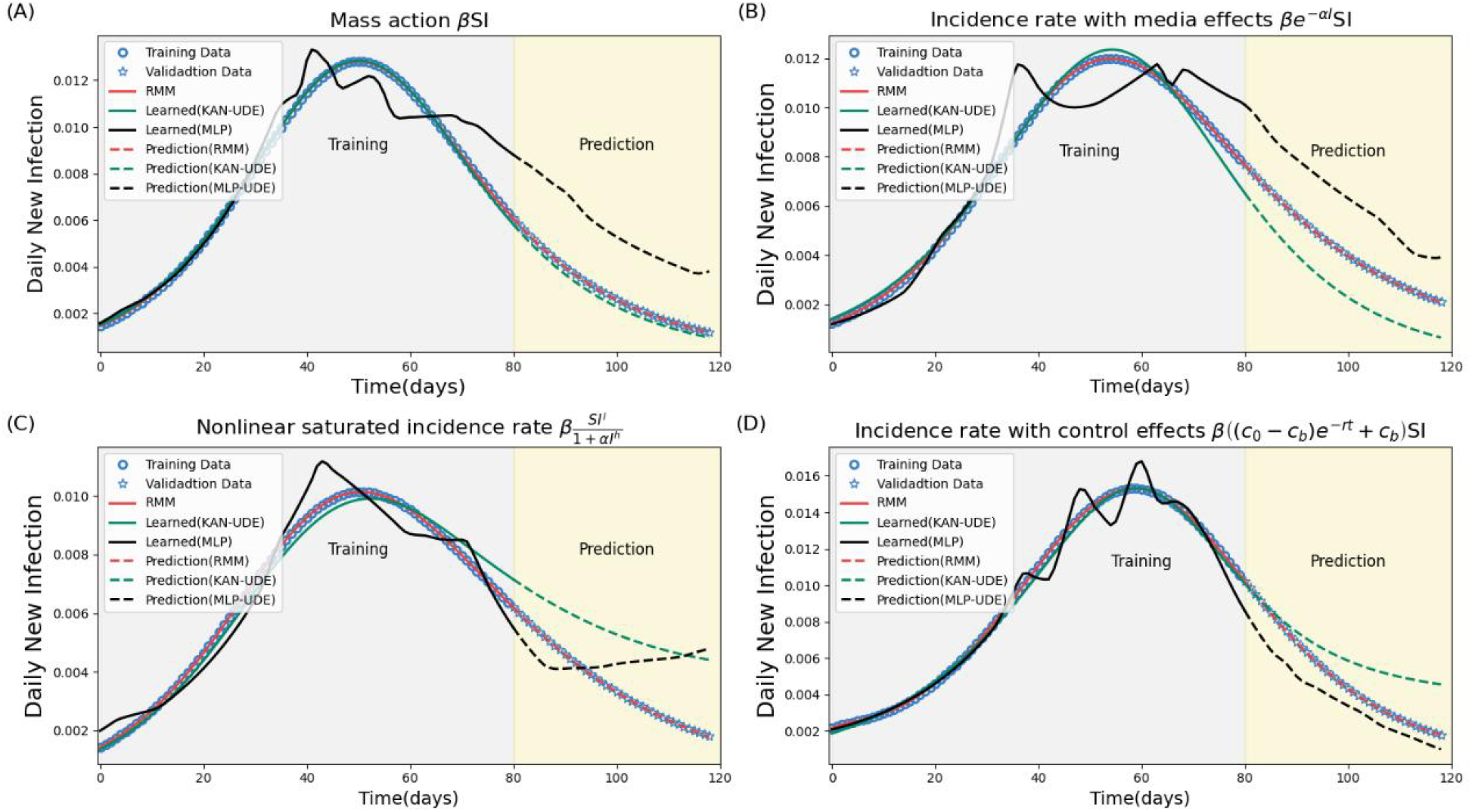
Epidemic prediction by KAN-UDE model, MLP-UDE models, and RMM by training the model with a subset of the time series data. Here the data from original period up to 80 day is used for training and the remaining data (validation data) for testing the prediction accuracy. The training process is stopped after 5,000 iterations.

### Robustness analysis

In this section, we use noisy data to train the KAN-UDE model, reconstruct the incidence rate function, and predict the epidemic trends of daily new infections. The main findings are presented in Fig. 9 and Fig. 10. These results are compared to those obtained from the original data, which is also plotted in the figures for convenience. As shown in Fig. 9, the KAN-UDE model can also effectively fit the noisy data, and well capture the epidemic trajectories. And the learned curves from the noisy data can also match the curves learned from the original data. However, the UDE model’s predictions show poor robustness, with a significant difference between the predicted curves generated by the KAN-UDE models trained on original versus noisy data. Despite this, the KAN-UDE model still accurately and robustly reconstructs the incidence rate functions across various types when trained with noisy data. Consequently, the RMMs learned from noisy data can also produce robust and highly accurate predictions, comparable to those obtained from RMMs trained on original time series data.

**Figure 9.**
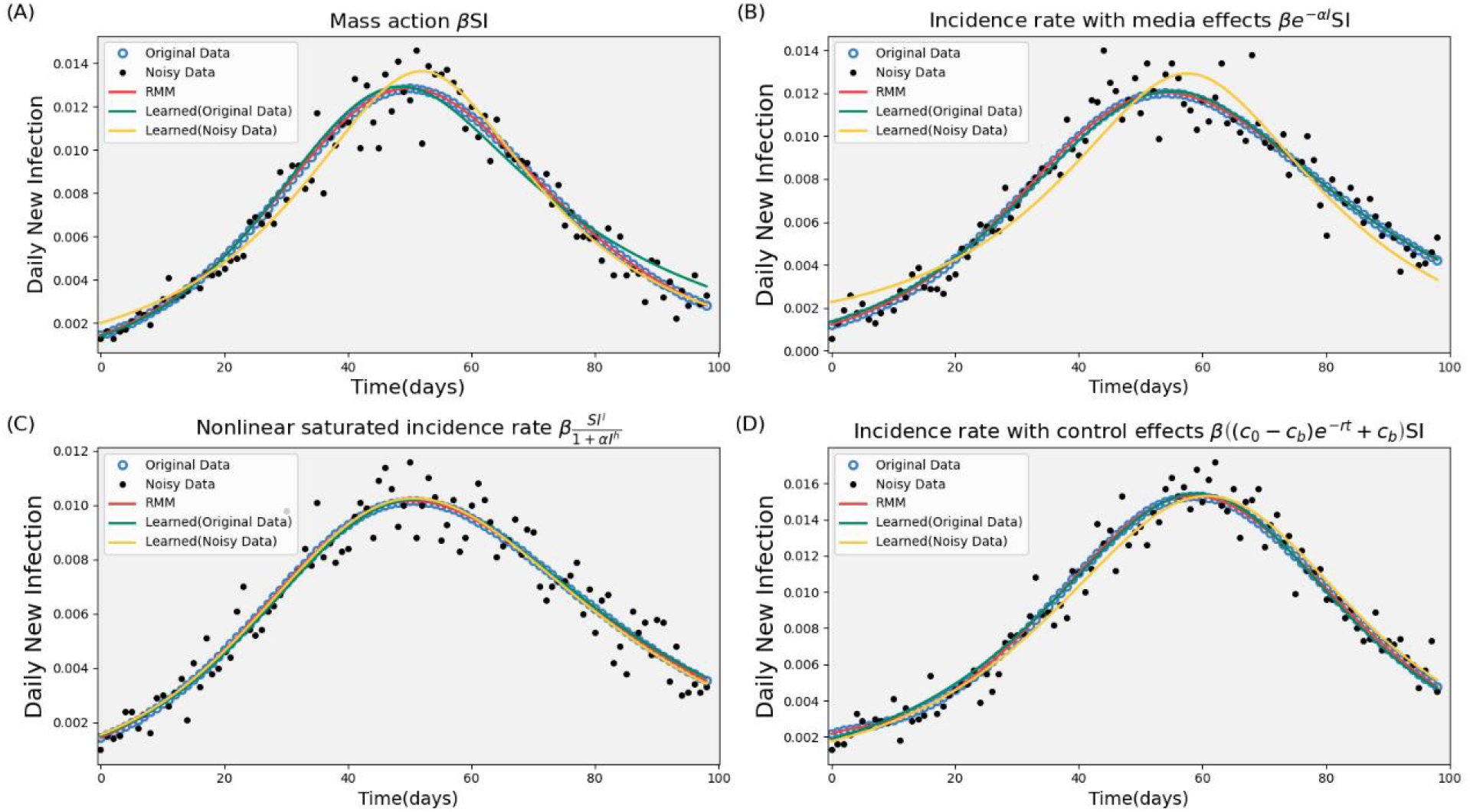
Training results on the noisy data in terms of the four types of incidence rate. The training process is stopped after 5,000 iterations. Here the RMMs are obtained by reconstructing the functions using the noisy data. For comparison, the training results on the original data are also displayed in the figures.

**Figure 10.**
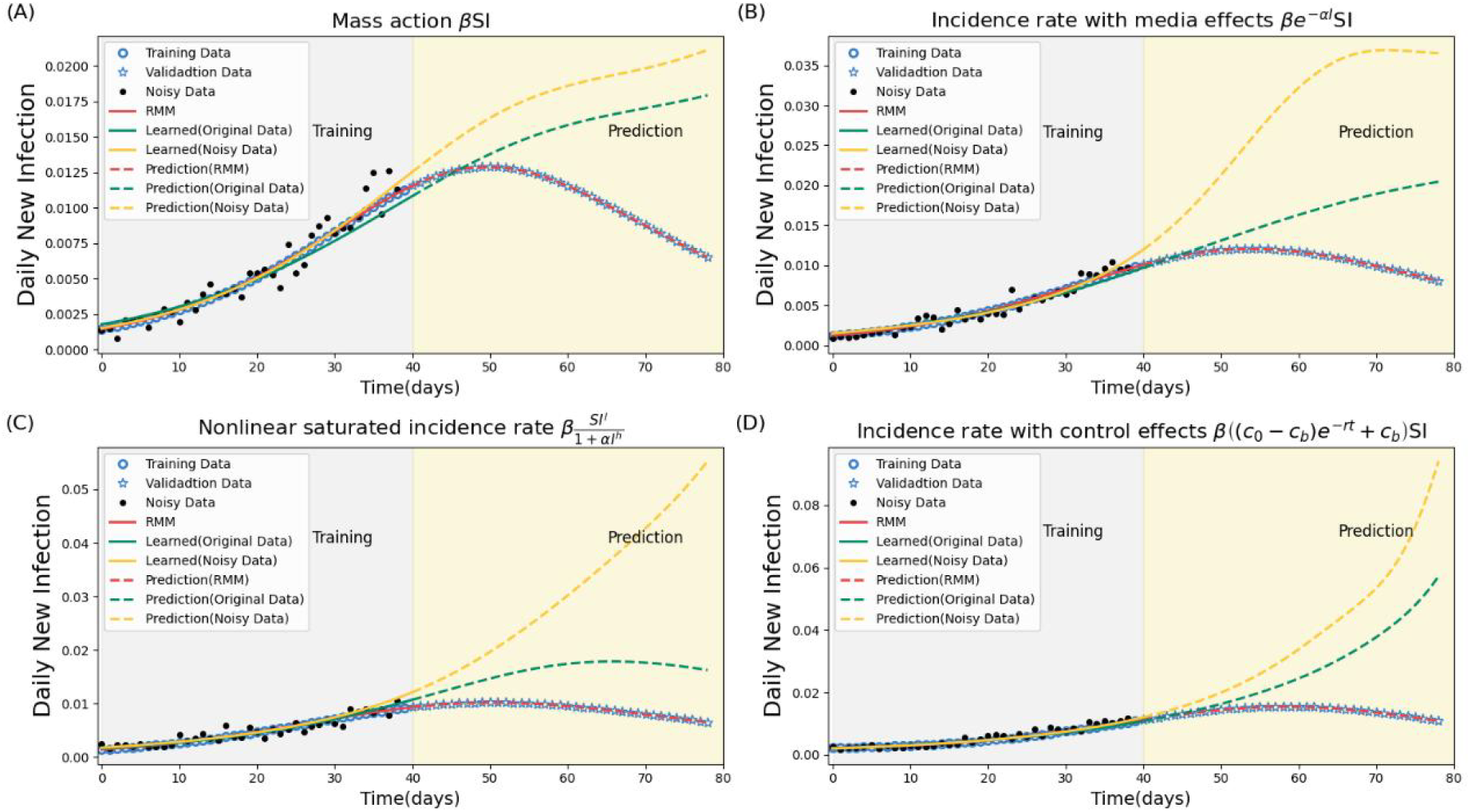
Epidemic predictions made by the UDE models trained on either noisy or original data. In this case, the data from the first 40 days is used for training, while the remaining data (validation data) is used to test prediction accuracy. The training process is stopped after 5,000 iterations.

## Discussion

Recently, an innovative neural network framework, Kolmogorov-Arnold Networks (KAN), has been proposed, where the Kolmogorov-Arnold representation theorem underpins the approach, proposing that any multivariate continuous function can be represented as a superposition of continuous functions of one variable. Several studies have already pioneered their attempts to replace the MLP-based neural network by KAN^24^, aim at testing the performance of KAN in learning process^25–28^. This study focuses on embedding KANs within mechanistic ODE model frameworks, creating a hybrid model that retains the physical interpretability of differential equations while enhancing the ability to reconstruct the mechanistic formulation of complex and nonlinear interactions that occur in many scientific fields.

As concluded in previous research^29^, biomedical studies primarily focus on two objectives: prediction and explanation. Using epidemic modeling in emerging infectious diseases as a case study, we established a modeling framework that couples efficient, interpretable, and robust deep learning approaches by leveraging the power of the Kolmogorov-Arnold Network (KAN). This framework is designed to learn mechanistic functions in epidemic models and to provide precise and robust predictions of epidemic trends, thereby offering new insights into the integration of mechanistic models with deep learning.

It is well known that the incidence rate and transmission rate are critical components of epidemic modeling, yet the form and parameter values of the incidence rate are often difficult to determine. Therefore, we focused on the incidence rate as the element to be learned and proposed the corresponding epidemic KAN-UDE and MLP-UDE models. By training these models with time series data of daily new infections or reported cases, we found that training of KAN-UDE models are much more efficient compared to the training of MLP-UDE models, both in terms of achieving higher accuracy after the same number of iterations and in requiring fewer iterations to reach the same level of accuracy. This supports the conclusion that KAN outperforms MLP in achieving higher accuracy.

To achieve interpretable deep learning for understanding the spread of infectious diseases, we reconstructed the incidence rate functions, represented by neural networks in the KAN-UDE models. We demonstrated that KAN accurately reconstruct these functions, closely mirroring the corresponding incidence rate of the mechanistic models used to generate the time series data. This approach helps to make the “black box” elements of coupling mechanistic models with deep learning more interpretable, resulting in a fully mechanistic model (referred to as RMM). Notably, even when training the model using only a subset of the time series data (specifically, data from before the turning point of an epidemic outbreak), KAN was able to robustly and precisely reconstruct the incidence rate function. This is critical, as we often have very limited data during the initial phase of epidemic outbreaks.

In addition to enhancing interpretability, reconstructing the mechanistic function is also crucial for achieving robust and precise predictions of epidemic trends. Given the inherent randomness in real data, we trained the model using noisy data and then reconstructed the incidence rate function, which matched exactly with the function learned from the original data. As a result, the RMM models trained on noisy data produced the same robust and precise long-term predictions of epidemic trends. In contrast, when predicting epidemic trends directly using the KAN-UDE models, the predictions based on the original data and the noisy data differed significantly, indicating poor robustness. Notably, the accuracy of long-term predictions by the KAN-UDE model was also substantially lower.

It is important to note that we also used time series data of daily reported cases for training, epidemic predictions, and robustness analysis, including training on noisy data, as shown in SI Figs. 2-8. We observed similar results in terms of the training accuracy of the KAN-UDE models, precise function reconstruction, and accurate epidemic predictions when compared to using time series data of daily new infections to train the models. This is crucial for data-driven approaches in epidemic analysis, as different epidemics may present varying types of data given the data accessibility.

As we mentioned in the introduction, dynamic systems are integral to many fields. Therefore, the modeling framework that couples KAN with differential equations should have wide-ranging applications across numerous domains. In addition to the mechanistic analysis and more accurately prediction for the spread of infectious diseases, thereby supporting public health decision-making, the potential applications of KAN-UDE modelling framework are vast and varied, extending beyond the examples mentioned here. In the field of financial engineering, it could model the dynamics of asset prices, providing more precise tools for risk management. Additionally, in meteorology, KAN-UDE models could enhance weather forecasting models, leading to better predictions of extreme weather events. Furthermore, in ecology, it could be employed to simulate population dynamics within ecosystems, offering theoretical support for environmental conservation and sustainable development. In summary, the application potential of KAN-UDE models is extensive, and its capabilities merit further exploration and development.

## Conclusion

In conclusion, using epidemic modeling of infectious diseases as a case study, this research demonstrated that KAN significantly improves learning accuracy and provides an interpretable deep learning approach for reconstructing mechanistic functions in complex dynamic systems. It also exhibits high robustness when trained on subsets of time series data or noisy data. Our approach paves the way for a new paradigm in modeling dynamical systems, where the integration of machine learning and traditional modeling techniques yields more accurate, interpretable, and robust predictions. This integration offers a flexible and powerful representation of a system’s physical processes, accommodating both known and unknown components. This work not only advances the field of computational modeling but also provides practical insights and tools for scientists and engineers addressing complex dynamic systems.

## Data Availability

All data produced in the present work are contained in the manuscript

## Author contributions

Conceptualization, KM, BT; validation and simulation, KM, XL; data curation, KM, XL, BT; writing original draft preparation, KM, BT; writing review and editing, NB, BT; Supervision: BT. All authors have read and agreed to the published version of the manuscript.

## Data accessibility

All the data used in this study are also available on request to the corresponding authors.

## Declare of interests

The authors declare no competing interests.

