## Supplementary information for "Integrating Kolmogorov-Arnold Networks with Ordinary Differential Equations for Efficient, Interpretable and Robust Deep Learning: A Case Study in the Epidemiology of Infectious Diseases"

**Definition of Training Accuracy.** We provide the definition of training accuracy for the time series data. Specifically, for each time point i, the distance between the simulated data $y_i$ and the projected value $\tilde{y}_i$ from the trained UDE models is defined as $d_i = |\tilde{y}_i - y_i|$. We consider the result as accurately trained for each point if the distance $d_i$ is less than a critical value $\delta$, which is set to 0.01 by default. Denote

$$c_i = \begin{cases} 1, & if\ d_i < \delta, \\ 0, & otherwise. \end{cases}$$

Then, $N_{accurate} = \sum_{i=1}^{n} c_i$ is the number of the time points with accurate training as defined above. The training accuracy is then calculated as the ratio of $N_{accurate}$ to the total number $N_{total}$ of time points in the time series, and is given by:

$$\text{Accuracy} = \frac{N_{accurate}}{N_{total}} \times 100\%.$$

### SI Figures

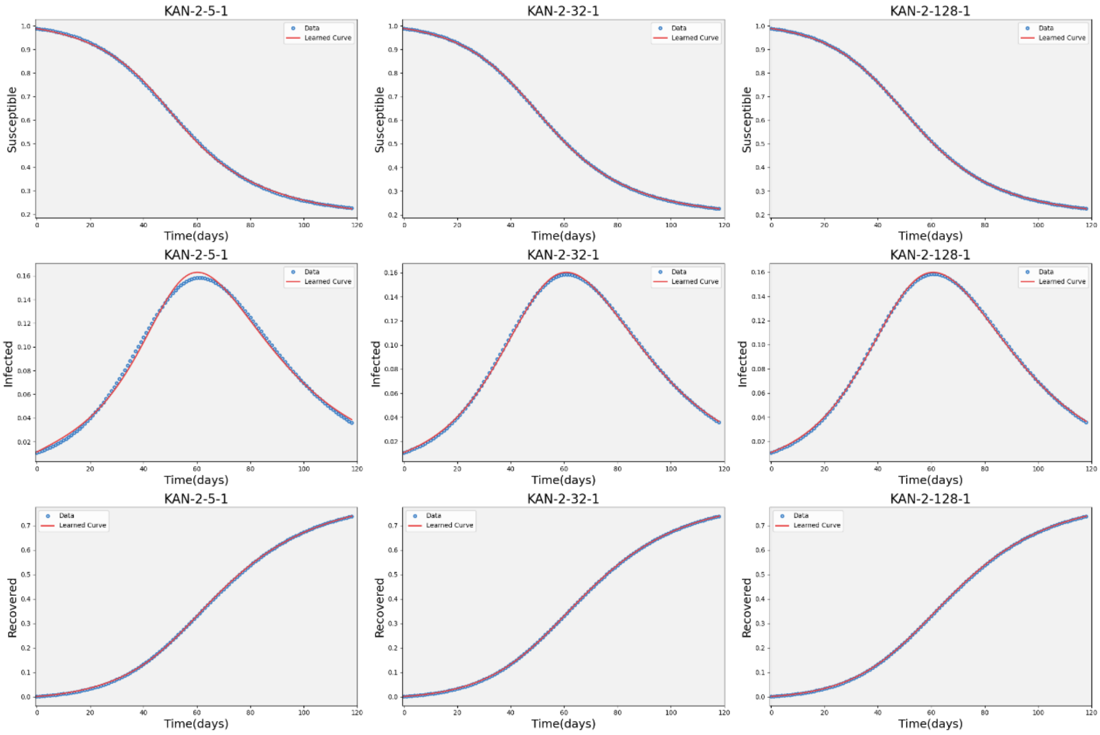


**SI Figure 1.** Solutions of trained KAN-UDE models with three types of network configuration as marked in the title of each sub-figure. Here, the model is trained using the time series data of daily new infections generated by the mechanistic model with the incidence rate of mass action. The data of the solutions of the three variables in model (4) are obtained when we generate the time series data of daily new infections by running model (4).

In what follows, we use the time series data of daily reported cases, which are also generated by the mechanistic models with various types of incidence rate listed in Table 1 in the main text, to repeat the key results of training the UDE models, conducting epidemic predictions and robustness analysis. The main observations are shown in SI Figs. 2-8. The methods are the same to those used in main text.

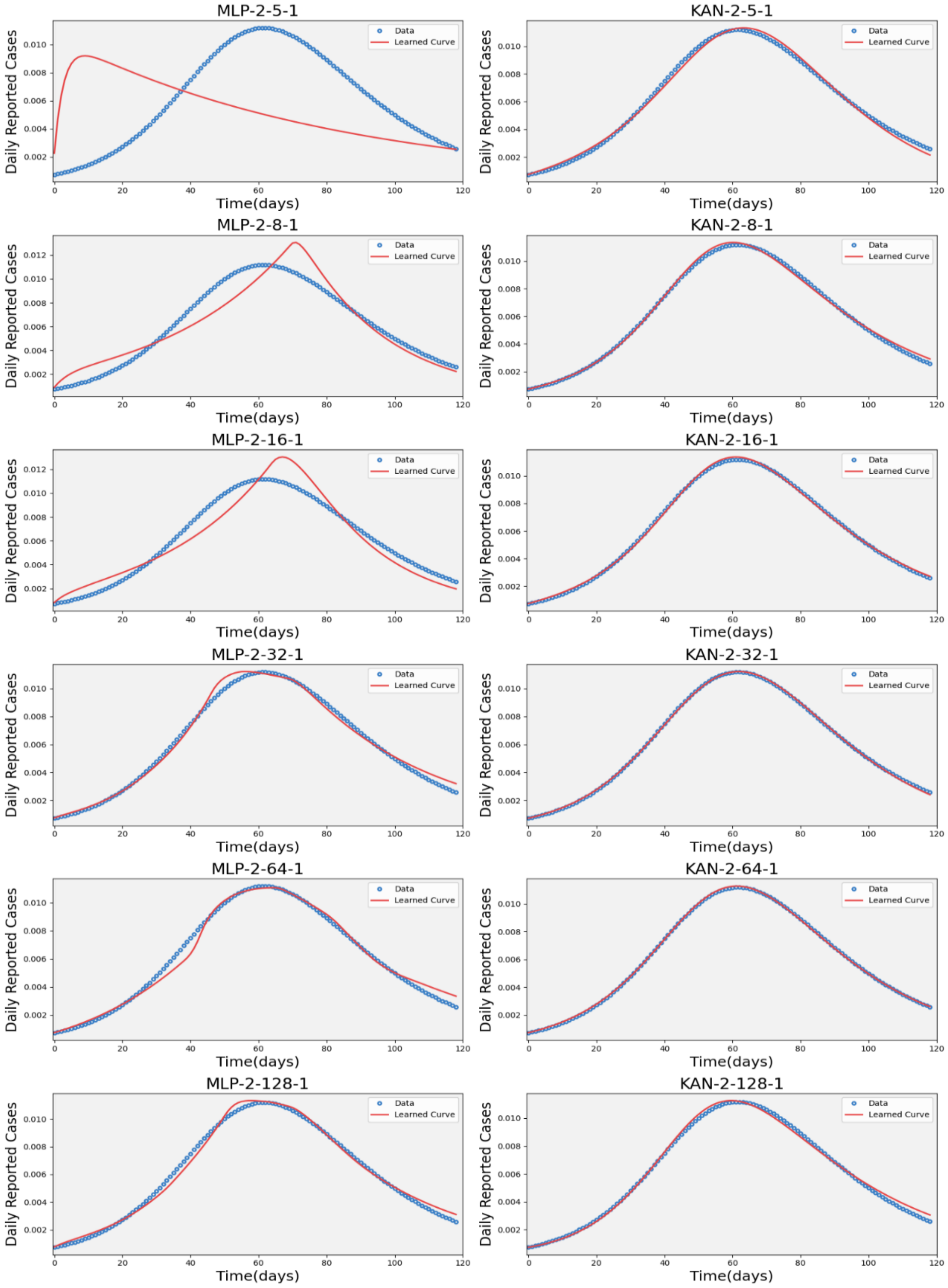


**SI Figure 2.** Training results of MLP-UDE model and KAN-UDE model, with six type of network configuration, on time series data of daily reported cases. Here the data is generated from the mechanistic model with the incidence rate of mass action. The training process is stopped after 5,000 iterations.

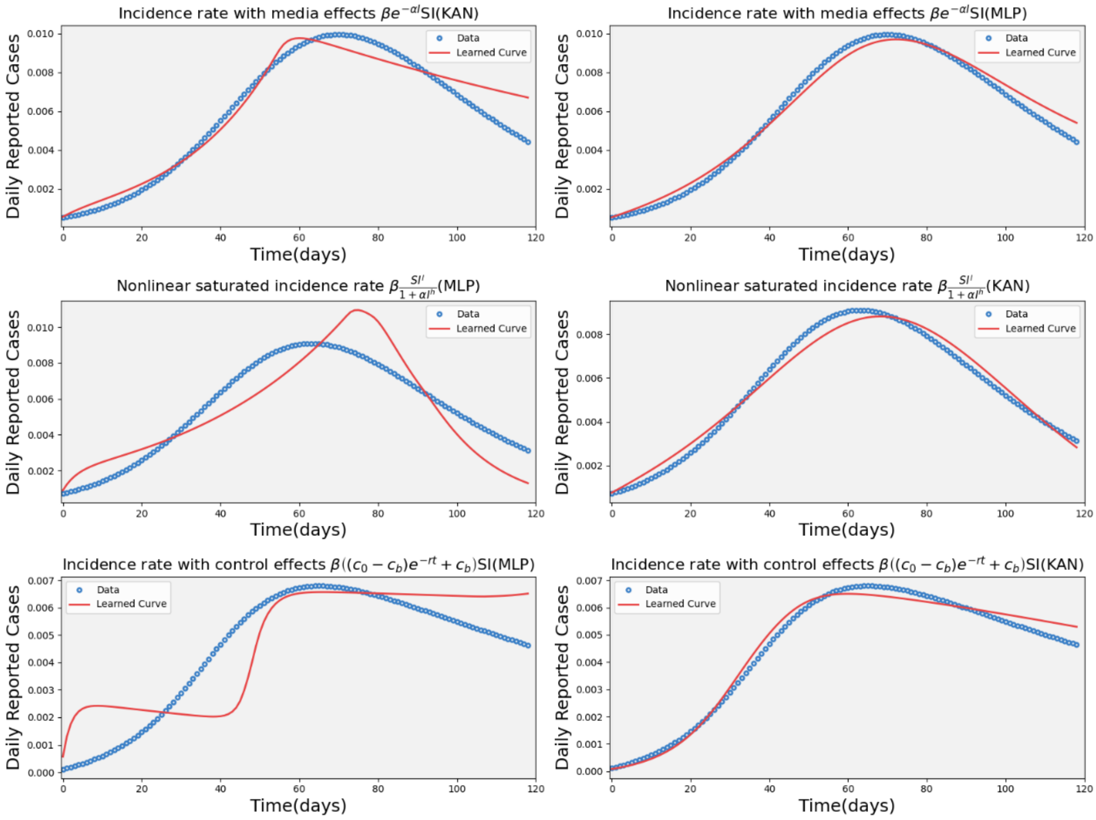


**SI Figure 3.** Training results of MLP-UDE model and KAN-UDE model on time series data of daily reported cases generated from the mechanistic model with the other three types of incidence rate, as marked in the title of each sub-figure. Here, the network configuration is fixed as 2-16-1. The training process is stopped after 5,000 iterations.

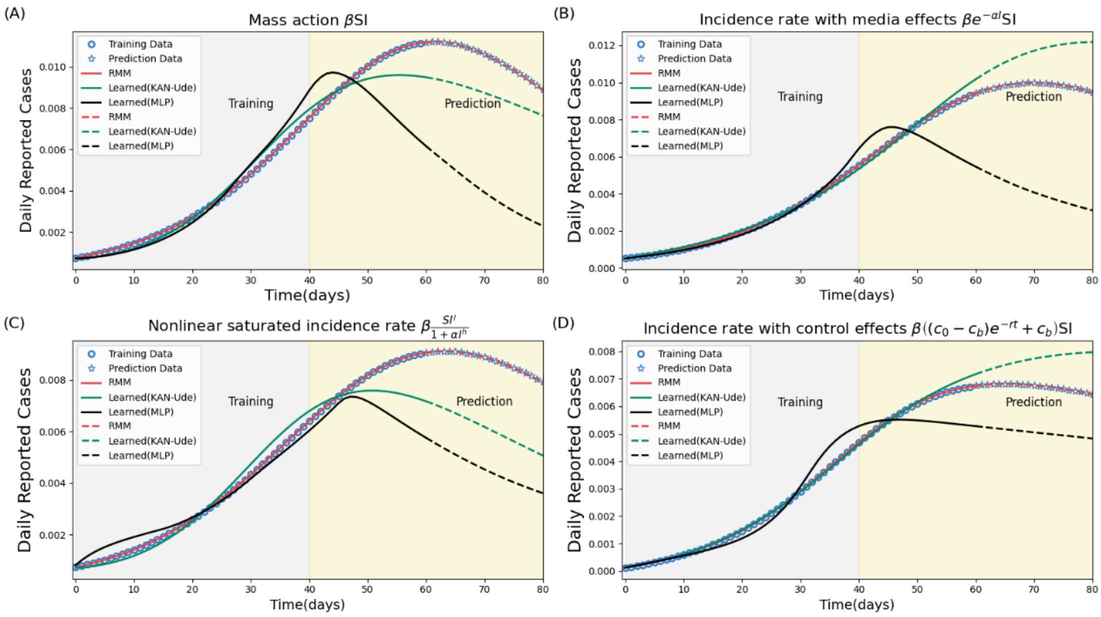


**SI Figure 4.** Epidemic prediction by KAN-UDE model, MLP-UDE models, and RMM by training the model with a subset of the time series data of daily reported cases. Here the data from original period up to 40 day is used for training and the remaining data (validation data) for testing the prediction accuracy. The training process is stopped after 5,000 iterations.

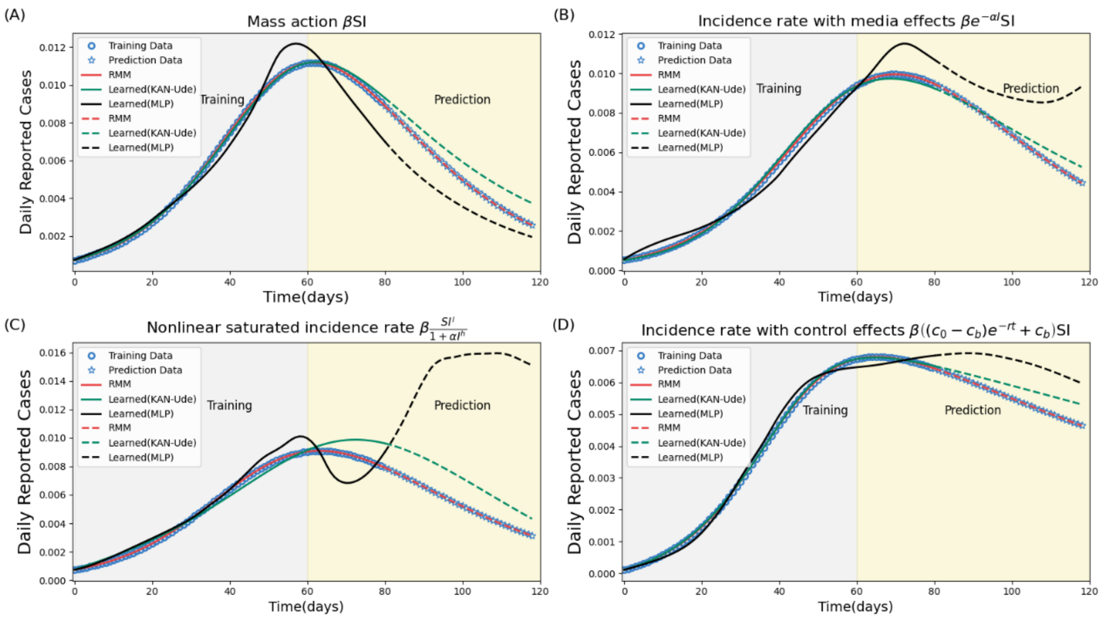


**SI Figure 5.** Epidemic prediction by KAN-UDE model, MLP-UDE models, and RMM by training the model with a subset of the time series data of daily reported data. Here the data from original period up to 60 day is used for training and the remaining data (validation data) for testing the prediction accuracy. The training process is stopped after 5,000 iterations.

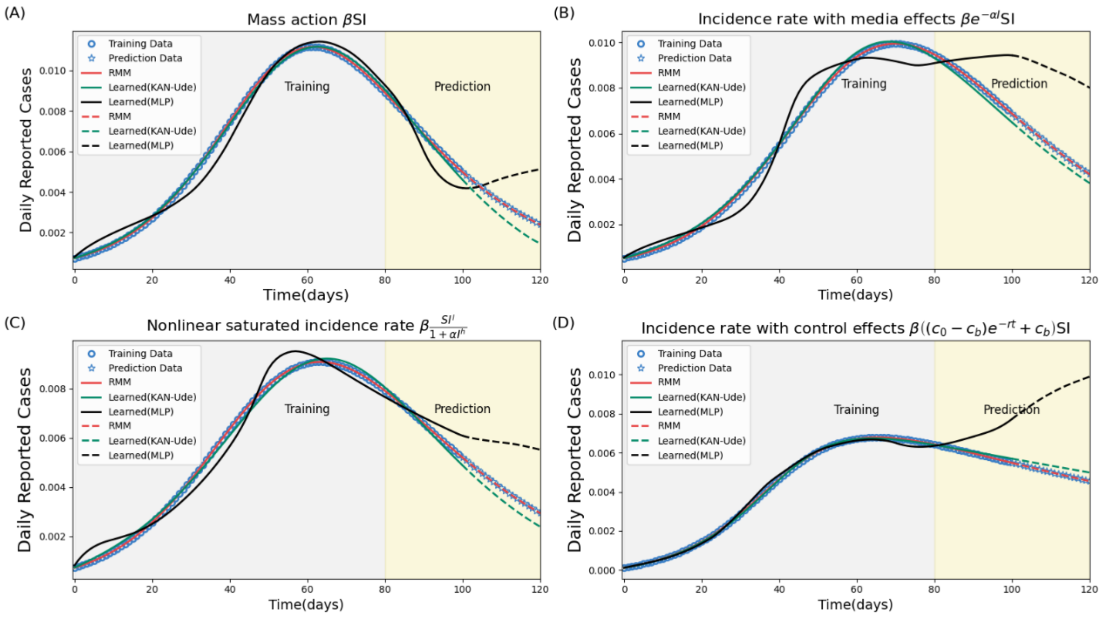


**SI Figure 6.** Epidemic prediction by KAN-UDE model, MLP-UDE models, and RMM by training the model with a subset of the time series data of daily reported cases. Here the data from original period up to 80 day is used for training and the remaining data (validation data) for testing the prediction accuracy. The training process is stopped after 5,000 iterations.

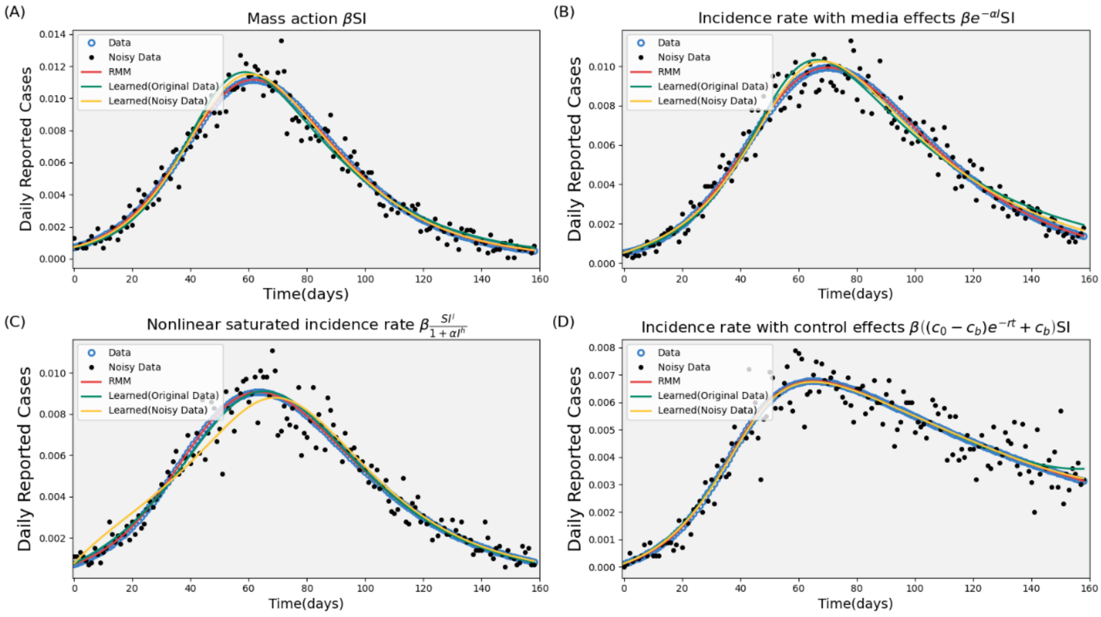


**SI Figure 7.** Training results on the noisy data of the time series data of daily reported cases generated by the mechanistic models with the four different types of incidence rate. The training process is stopped after 5,000 iterations. Here, we also displayed the learned curves from the original data for comparison.

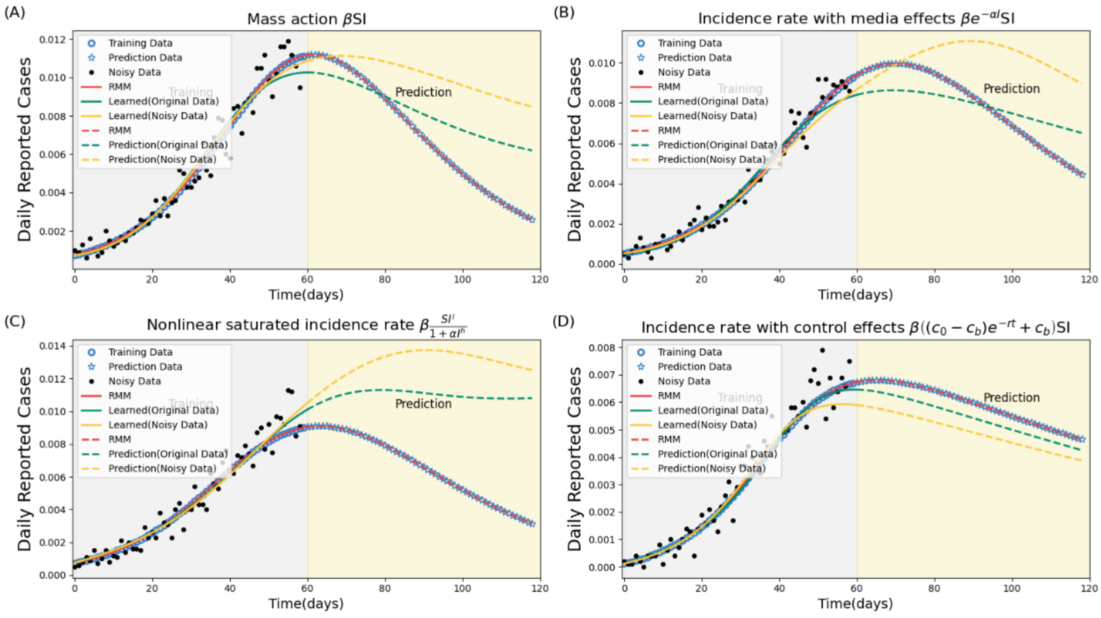


**SI Figure 8.** Epidemic predictions by the UDE models learned from the noisy data or the original data. Here the time series data of daily reported cases from the original period up to 40 day is used for training and the remaining data (validation data) for testing the prediction accuracy. The training process is stopped after 5,000 iterations.
